# Evaluating the Performance of Predicting *Plasmodium vivax* Infection Risk Using Serological Markers in Patients with *Plasmodium falciparum* Malaria

**DOI:** 10.1101/2025.06.26.25330308

**Authors:** Maulina Hafidzah, Tamiru Shibiru Degaga, Michael Christian, Mohammad Shafiul Alam, Mohammad Sharif Hossain, J Kevin Baird, Inge Sutano, Lauren Smith, Dionne Argyropoulos, Aisah R Amelia, Rintis Noviyanti, Ric N Price, Ivo Mueller, Kamala Thriemer, Rhea J Longley

## Abstract

**Background:** *Plasmodium vivax* presents a significant obstacle to malaria elimination due to its capacity to form dormant liver-stage hypnozoites that can cause relapses. Universal radical cure, which administers hypnozoite-targeting treatment to patients with *P. falciparum* malaria living in co-endemic areas, has potential to reduce *P. vivax* relapses. However, its implementation is hindered by the lack of a diagnostic tool for detecting hypnozoite carriage.

**Methods:** This study evaluated the performance of *P. vivax* serological exposure markers (SEMs) as a screening tool for predicting individuals with clinical *P. falciparum* infections at risk of *P. vivax* relapse over the following 63 days. Analysis was performed using samples from participants in the PRIMA study from Ethiopia, Indonesia and Bangladesh (NCT03916003). An existing random forest classification model was used and evaluated for sensitivity and specificity, then further optimized by excluding potentially cross-reactive antigens and re-training a study-specific model.

**Results:** IgG antibodies were measured in 244 *P. falciparum* participant samples, of which 22 had a vivax episode during follow-up. SEMs showed high sensitivity (82%) but low specificity (27%), likely due to sustained antibody responses in moderate-to-high transmission areas. Excluding potentially cross-reactive antigens yielded minimal improvements. A study-specific algorithm increased specificity to 68%, but with a corresponding drop in sensitivity to 68%.

**Discussion:** Although SEMs showed limited suitability for guiding radical cure in high-transmission settings, they demonstrate promising potential in low-transmission areas where their performance may be more impactful. SEMs could also play a valuable role in building community trust and acceptance of universal radical cure when supported by strong communication and implementation strategies. Further research is needed to evaluate SEM-based approaches across varying transmission contexts and their potential role in supporting malaria elimination efforts.

## INTRODUCTION

Over the last two decades, significant progress has been made in reducing the global burden of malaria through expanded access to tools preventing infection and treating parasitaemic individuals.^1^ Despite these advances, *Plasmodium vivax* remains a significant challenge for malaria elimination efforts. Whilst many malaria control efforts have focused on *Plasmodium falciparum* due to its high associated mortality, *P. vivax* contributes to a substantial proportion of global malaria cases and imposes a greater long-term burden through repeated relapses and hospitalizations.^2^ Unlike most *Plasmodium* species, *P. vivax* can form liver-stage hypnozoites that can reactivate and trigger recurrent infections, leading to clinical relapses unless treated with specific liver-stage-targeting drugs.^3^ Effective clearance of hypnozoites requires the use of 8-aminoquinolines, such as primaquine or tafenoquine.^4^ However, this treatment requires glucose-6-phosphate dehydrogenase (G6PD) testing to minimise adverse events, such as haemolytic anaemia, in patients with G6PD deficiency.^4^ Additionally, control efforts are confounded by the lack of commercial diagnostics to identify individuals with hypnozoites^5^, occurring in the absence of patent blood-stage infection. With approximately one-third of the world’s population at risk of *P. vivax* infection^6^, there is an urgent need for improved diagnostic and treatment strategies.

The PRIMA study was a multicentre, open-label, randomized controlled trial conducted in Bangladesh, Indonesia, and Ethiopia, to evaluate primaquine radical cure of patients with *P. falciparum* malaria as a novel approach to reduce subsequent *P. vivax* relapses.^7^ This intervention was designed to address the high incidence of *P. vivax* parasitemia following treatment for *P. falciparum* malaria.^8,9^ The study demonstrated that patients treated with a high-dose primaquine regimen (1.0 mg/kg per day for 7 days) had fivefold lower risk of *P. vivax* recurrence compared to those treated with standard care (0.25 mg/kg single-dose primaquine to treat *P. falciparum* gametocytes).^7^ These findings highlighted the potential to expand the indication for radical cure to patients who present with *P. falciparum* malaria, an approach termed “universal radical cure”. However, the acceptability of universal radical cure is limited by concerns of exposing patients who are not infected with hypnozoites to the risk of primaquine induced haemolysis and thus the need to expand G6PD testing.^10^ Better strategies for targeting patients suitable for universal radical cure are needed.

*P. vivax* serological exposure markers (SEMs) can be used to identify individuals previously exposed to infection with *P. vivax*.^11^ These markers rely on detecting antibodies that indicate a history of recent *P. vivax* infection, which may correlate with an elevated risk of relapse due to the presence of arrested liver-stage hypnozoites.^11^ The *P. vivax* SEMs currently have 80% sensitivity and 80% specificity for accurately detecting recent *P. vivax* blood-stage infections in the prior 9-months^11^, and have been validated in multiple geographic regions including Asia (Thailand^11,12^, Cambodia^13^, Indonesia^14^), the Pacific (Solomon Islands^11^), and South America (Brazil^11^, Peru^15^). Nearly all strains of *P. vivax* exhibit a primary relapse within the first 9-months after their initial infection^16^, hence *P. vivax* SEMs have potential to act as indirect biomarkers for the risk of relapse. The ability to quantify the patients at risk of relapse led to the World Health Organization (WHO) releasing the first Preferred Product Characteristics (PPCs) for diagnosing relapse risk. The PPCs define various use cases for such a diagnostic test, including guiding radical cure, with use case 1b specifying acute case management aimed at preventing *P. vivax* relapses following *P. falciparum* infection.^17^ The diagnostic algorithm has not previously been evaludated in *P. falciparum* patient samples for this use-case.

The integration of *P. vivax* SEMs into radical cure approaches offers a potential advance in classifying individuals at higher risk of *P. vivax* relapse following a *P. falciparum* infection, who should be targeted for universal radical cure. The SEMs and the associated predictive algorithm could be used by healthcare professionals in countries co-endemic for both *P. falciparum* and *P. vivax* to guide use of radical cure treatment in individuals with clinically diagnosed *P. falciparum* infection. This approach could improve treatment outcomes, and minimize unnecessary primaquine use, thus reducing the risk of drug-induced adverse events. To evaluate the utility of SEMs in this context, we conducted a retrospective study using plasma samples collected from the patients enrolled into the PRIMA study, who were not treated with primaquine and thus remained at risk of relapse. Plasma samples were analysed for antibodies against the *P. vivax* SEMs using a multiplexed bead-based assay and data were analysed with a random forest algorithm, developed and validated based on prior data^11^, to evaluate the ability of the SEMs to classify *P. falciparum* patients at risk of future *P. vivax* relapse. Modified versions of the existing prediction model were also developed to better match the epidemiology of the tested setting.

## METHODS

### Ethical approval

Samples for this study were obtained from clinical trials conducted in Bangladesh, Ethiopia, and Indonesia as part of the PRIMA study run from August 18, 2019 to March 14, 2022. Ethical approval for these trials was granted by the NT Health and Menzies School of Health Research Human Research Ethics Committee (HREC), as well as by the independent review boards at each participating site.^7^ The clinical trial is registered on ClinicalTrials.gov under the identifier NCT03916003. Written informed consent was obtained from participants or their guardians prior to enrolment, and written assent was additionally secured from participants aged 11–18 years. Use of samples in Melbourne for serological analysis was approved by the WEHI HREC (23/35). The PRIMA study protocol was published previously (https://trialsjournal.biomedcentral.com/articles/10.1186/s13063-022-06364-z) and no separate protocol was made for the current study. No patients or public were involved in any aspect of the study design, conduct, reporting, interpretation, or dissemination.

### Study Samples: *P. falciparum* patients and positive controls

Day 0 plasma samples collected from *P. falciparum* patients enrolled in the PRIMA clinical trial, conducted in Bangladesh (one health clinic in Alikadam), Ethiopia (one health clinic in Arba Minch), and Indonesia (three health clinics, one each in Mangili, Waijeli, and Tanaraing), were utilised for this study. The PRIMA study included patients with uncomplicated *P. falciparum* mono infection who had experienced fever or a history of fever within 48 hours prior to their clinic visit. Eligibility criteria required participants to have a G6PD activity level of at least 70% of the adjusted male median for the study population, determined using the STANDARD G6PD test (SD Biosensor, Gyeonggi-do, South Korea). Exclusion criteria varied slightly by location: infants under 1 year of age were excluded in Bangladesh and Indonesia, while participants under 18 years were excluded in Ethiopia. Patients were also excluded if they exhibited signs or symptoms of severe malaria, had a haemoglobin concentration below 8 g/dL, were pregnant or breastfeeding, had a known hypersensitivity to the study drugs, were regularly using medications with haemolytic potential, or had received a blood transfusion within the previous four months.

Participants were actively monitored throughout the follow-up period, which extended from day 7 to day 63, with scheduled weekly visits to the health facility. Prior to this, during days 1 to 6, patients attended daily clinic visits for directly supervised administration of schizontocidal and primaquine treatment. In cases where artemether–lumefantrine was prescribed, only the morning dose was administered under supervision, while the evening dose was taken independently at home. In the standard care arm, patients received a single oral dose of primaquine (0.25 mg/kg) in conjunction with their schizontocidal treatment, as per national treatment guidelines. This dose was supervised in the same manner as for those in the intervention arm. Participants were advised to seek medical attention at the study centre should they develop any symptoms outside scheduled visits. Those who missed follow-up appointments were contacted via telephone or in person and encouraged to return for clinical assessment. In cases where participants were unable to attend, study staff conducted home visits. The study cohort consisted of 500 participants across the three study sites—99 in Indonesia, 332 in Ethiopia, and 49 in Bangladesh. However, only 474 participants were included in the final data analysis. This reduction was due to the exclusion of some patients who were not part of the original PRIMA paper analysis. These patients were excluded because they did not complete the 7-day PQ treatment, had severe malaria, very low haemoglobin levels, or were found not to have a *P. falciparum* mono-infection at the start. To maintain consistency with the original study criteria, only participants included in the trial data analysis were considered for the final analysis in this paper. For evaluating the predicting algorithm, and for training the new dataset specific algorithm, only control arm samples were utilised (n=244), whilst the whole 474 were used to analyse impact of various epidemiological and demographic factors on antibody levels.

Plasma samples from *P. vivax* positive controls from Papua New Guinea were also utilized as standard references for MAGPIX measurements.^18^ Additionally, negative control samples were obtained from a previous study, consisting of malaria-naïve individuals from Bangkok, Rio de Janeiro, and Melbourne^11^, which were used for comparison in the analysis.

### P. vivax proteins

The preparation of *P. vivax* protein-coupled beads followed established protocols.^18^ Briefly, BioPlex Pro-Magnetic COOH beads were initially sonicated and vortexed, and 50 μL of the microspheres were washed in MilliQ-H2O. For bead activation, 100 mM monobasic sodium phosphate was added to the beads, followed by 10 μL of hydroxysulfosuccinimide (sulfo-NHS) and 10 μL of N-Ethyl-N′-(3-dimethyl aminopropyl) carbodiimide hydrochloride (EDC) at 50 mg/mL. The beads were incubated in the dark on a rotating platform for 20 minutes. After activation, the beads underwent three washes with 1x phosphate-buffered saline (PBS) at pH 7.4. They were then resuspended in 1x PBS along with protein, with the optimized protein volume per antigen detailed in Table S1. The total volume of protein and PBS used was 250 μL. The beads were incubated overnight at 4 °C then washed three times with PBS-TBN (containing 0.1% bovine serum albumin (BSA), 0.02% Tween-20, and 0.05% azide, pH 7.4). After washing, the beads were resuspended in 112.5 μL of PBS-TBN. These stabilized beads were stored at 4 °C until further use.

The proteins used were selected from a previously established *P. vivax* serology panel, chosen for their ability to elicit strong antibody responses and classify individuals as recently exposed to *P. vivax* parasites within the preceding nine months, as described in earlier studies.^11^

### Total IgG antibody assay

IgG antibody binding was evaluated using the Luminex MAGPIX instrument, as previously described.^18^ A sample plate layout was prepared for the analysis, utilizing a total of seven 96-well plates to process all samples. In each well of the 96-well plate, 50 μL of *P. vivax* protein-coupled beads were combined with 50 μL of diluted plasma (1:100) and incubated for 30 minutes. After this, the beads were incubated with anti-human IgG phycoerythrin (1:100) for 15 minutes. The resulting bead complexes, comprising *P. vivax* protein, plasma sample, and anti-human IgG phycoerythrin, were then quantified using the Luminex MAGPIX system. To ensure the quality of the coupled beads, a log-linear standard curve was included on each plate. This curve, generated using hyperimmune plasma from Papua New Guinea, served as the assay standard. Additionally, two blank wells were included on each plate to confirm the reliability of the Luminex MAGPIX assay. All serology data from this study is available within Data S1.

### Plate quality control, plate standardisation and relative antibody unit conversion

The raw MFI values generated by the Luminex MAGPIX assay were converted to relative antibody units (RAU) using a protein-specific standard curve, as previously reported.^11^ This conversion was performed with a 5-parameter logistic regression model using the R ShinyApp developed by Dr. Shazia Ruybal-Pesántez (https://shaziaruybal.shinyapps.io/covidClassifyR/). As part of the quality control process, only wells containing more than 15 beads were considered valid, and blank wells were required to have MFI values below 50 to ensure the reliability of the assay results.

### Serology classification analysis

The outcome measured (that we were aiming to predict) was vivax episodes. These were detected throughout the 63-day follow-up by monitoring for malaria symptoms weekly, with detection via microscopy. This data was already available from the main PRIMA trial. In the current study, laboratory operators and statisticians were not blinded to the outcome. The antibody data (RAU from the RShiny app) generated in this study was were used to classify samples through a Random Forest algorithm, which determined seropositivity (indicative of *P. vivax* infection within the past nine months) or seronegativity (no *P. vivax* infection in the same period). Only antibody levels were used as predictors. Model outputs per individual therefore were a binary classification of recently exposed (seropositive) or not recently exposed (seronegative). This method uses multiple decision trees, with the final classification determined by a majority vote across all trees. Adjustments to the voting threshold allowed refinement of sensitivity and specificity parameters. The algorithm was previously trained using data from longitudinal cohort studies conducted in Thailand, Brazil, and the Solomon Islands^11^, utilizing a panel of eight *P. vivax* proteins previously identified as robust markers of recent exposure. These settings were considered low transmission. Whilst it is difficult to compare malaria transmission across countries and studies, we consider the Thai, Brazil and Solomon Islands cohorts to have lower transmission than the PRIMA study settings. Code is available at github.com/dionnecargy/PvSeroApp. A modified version of the algorithm, tailored to this dataset, was also implemented. This used the same available antibody data as predictors. As training and testing a random forest algorithm on the same dataset may lead to overfitting, we used 10-fold cross validation with five repeats to assess the classification performance. In this process, we train a random forest on 90% of the data, and test on the held out 10% to assess classification performance. This is repeated 10 times to use the whole dataset, and we repeat this process five times. Code is available at github.com/Longley-Lab/PRIMA. Various algorithms were utilised in this study, as outlined in Table S1. The classification performance of all algorithms were assessed through a confusion matrix (as shown in Table S2) with sensitivity, specificity, positive predictive value (PPV), and negative predictive value (NPV) based on the random forest algorithm with maximised threshold (maximum sensitivity and specificity) calculated as follows:

- 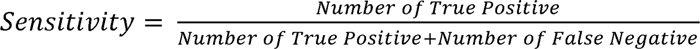, representing the proportion of true positives among seropositive classifications.
- 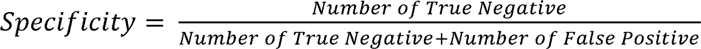, representing the proportion of true negatives among seronegative classifications.
- 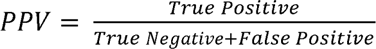, reflecting the likelihood of relapse in seropositive individuals.
- 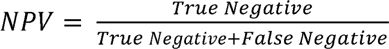, the likelihood of no relapse in seronegative individuals.

Performance was evaluated using all available data and at a country-specific level, with both results reported. When comparing models, all above values were considered as well as the area under the curve (AUC) of receiver operator characteristic (ROC) curves.

### Statistical Analysis

Statistical analyses were performed using RStudio (version 2024.04.0) and GraphPad PRISM (version 10.3.0). Differences in RAU between study sites and age groups were analysed using the Kruskal-Wallis test followed by Dunn’s post-hoc test, while comparisons between genders were assessed using the Mann-Whitney U test. These helped consider potential impacts on antibody levels, which in turn could influence classification performance (model fairness). ROC curves were generated for visualization, and the AUC was used as a summary measure of performance. Additionally, Kaplan-Meier curves were analysed to assess time-to-relapse data, with relevant statistical comparisons performed to evaluate differences between groups. A p-value of <0.05 was considered statistically significant. No formal sample size calculation was performed as all available data were used.

## RESULTS

### High sensitivity, but limited specificity, of *P. vivax* SEMs when applied to *P. falciparum*

### patient samples

The study assessed the application of *P. vivax* SEMs to evaluate relapse risk in *P. falciparum* patients. The primary goal was to determine whether *P. vivax* SEMs could reliably identify patients with *P. falciparum* malaria at greatest risk of *P. vivax* relapse, to facilitate targeted universal radical cure. IgG antibody levels were measured against a panel of 8 *P. vivax* antigens, and a previously trained random forest classification algorithm applied to the dataset to define each individual as recently exposed to *P. vivax* and at risk of relapse, or not (Figure 1A). Of the 244 individuals assayed, 181 were classified as seropositive and 63 as seronegative (Figure 1B). At the maximised random forest votes threshold, the algorithm had high sensitivity, correctly classifying 81.8% (18/22) *P. falciparum* individuals who presented with *P. vivax* parasitaemia during follow up, but poor specificity at 26.6% (59/222). There was a high rate of false positives, with many individuals classified as seropositive who did not have a subsequent *P. vivax* episode within 63 days (Figure 1B). Kaplan-Meier analysis (time-to-event) demonstrated that the episode-free probability of *P. vivax* remained high for both groups (seropositive or seronegative) throughout the study (Figure 1C), with minimal decline after 30 days. By day 60, the probability of patients being *P. vivax*-free was 91.8% for seronegative and 89.7% seropositive groups. Seronegative patients experienced a slightly steeper decline between days 30 and 40, but overall, there was no significant difference between the two groups (log-rank test, p=0.49). We hypothesized that the poor specificity could be due to antibody cross-reactivity with *P. falciparum*, in which *P. falciparum* parasitaemia boosted antibody response to *P. vivax* proteins, although this effect was not observed in the initial validation study of the *P. vivax* SEMs.^11^ Importantly, the negative predictive value (NPV) was high (93.7% (59/63), Figure 1B), with the majority of seronegative individuals having no detected vivax episodes during the follow-up period.

**Figure 1.**
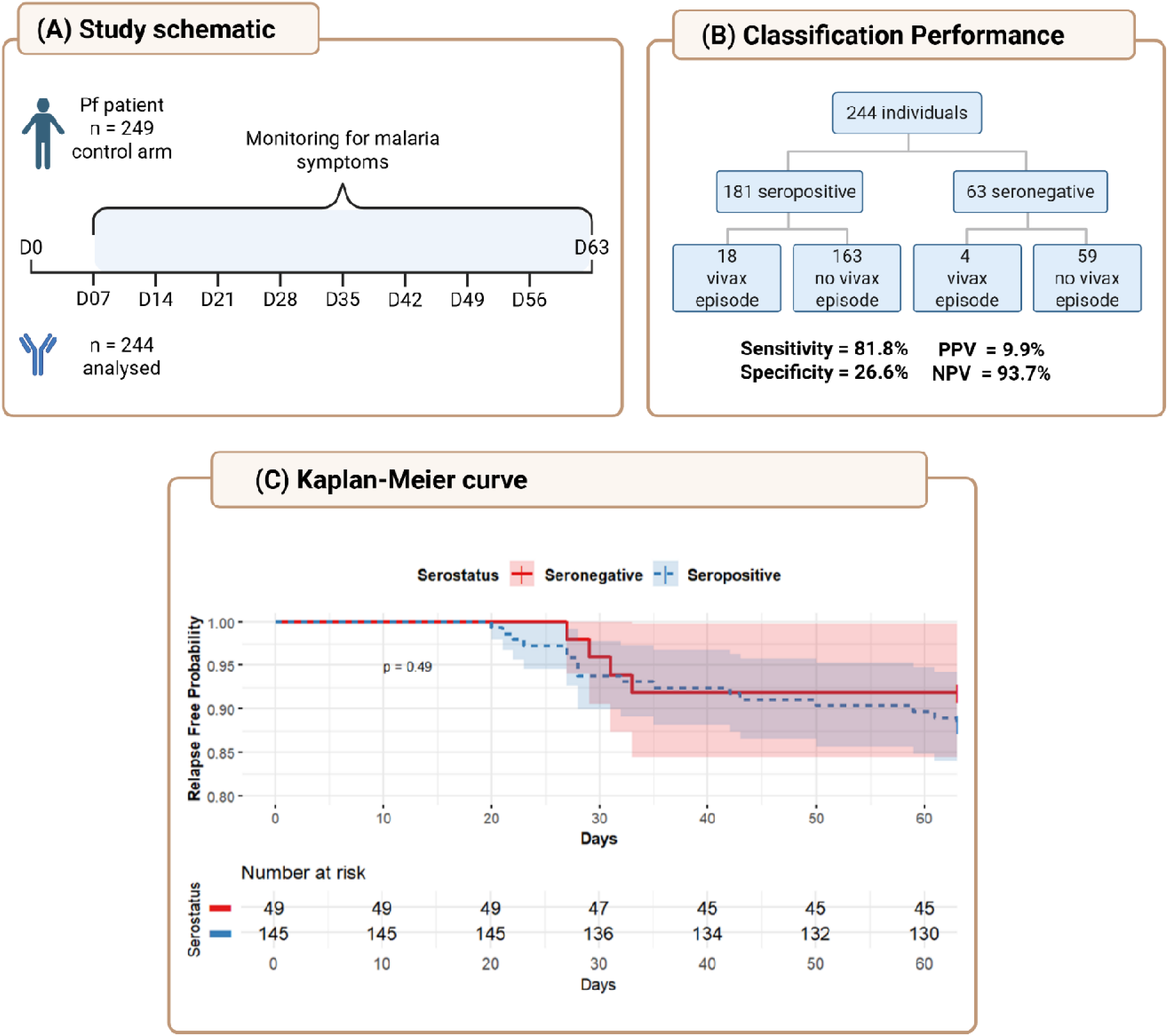
*P. vivax* SEM classification accuracy for predicting *P. falciparum* patients at risk of future vivax episode/s. **A)** Study schematic. **B)** Serostatus results in PRIMA control arm patients, and subsequent vivax episode outcome. Sensitivity, specificity, PPV and NPV are presented, calculated as detailed in the STAR Methods. Results split via country are provided in Figure S2. **C)** Kaplan-Meier analysis of time to *P. vivax* episode, with *P. falciparum* patients split via serostatus.

### Limited evidence of antibody cross-reactivity contributing to poor classification performance

All eight *P. vivax* proteins have low levels of sequence identity with their *P. falciparum* orthologs (<50%, Table S3), and we have demonstrated previously significant (>10-fold) cross-reactivity between *P. vivax* and *P. knowlesi* when sequence identity was >65%^19^. To address the potential impact of the *P. falciparum* infection boosting antibody levels to the *P. vivax* SEMs, we used a modified algorithm that excluded *P. vivax* proteins with known temporary increases in IgG levels 7-days post-clinical *P. falciparum*: RAMA, RBP2b, PTEX-150, and PvEBP (Wu *et al* in preparation). These proteins were excluded in the “Longitudinal Low Pf” algorithm, trained on existing data from the published longitudinal cohort studies.^11^ A second version, “Longitudinal Low Pf with RBP2b,” was also tested, given the importance of RBP2b in the classification.^11^ Excluding potentially cross-reactive proteins improved sensitivity from 81.8% (18/22) to 90.9% (20/22), but reduced specificity from 26.6% (59/222) to 14.0% (31/222). Including RBP2b increased specificity slightly to 31.1% (61/222) suggesting that excluding potential cross-reactive proteins did not significantly improve classification performance for the PRIMA dataset (Table 1).

**Table 1.**
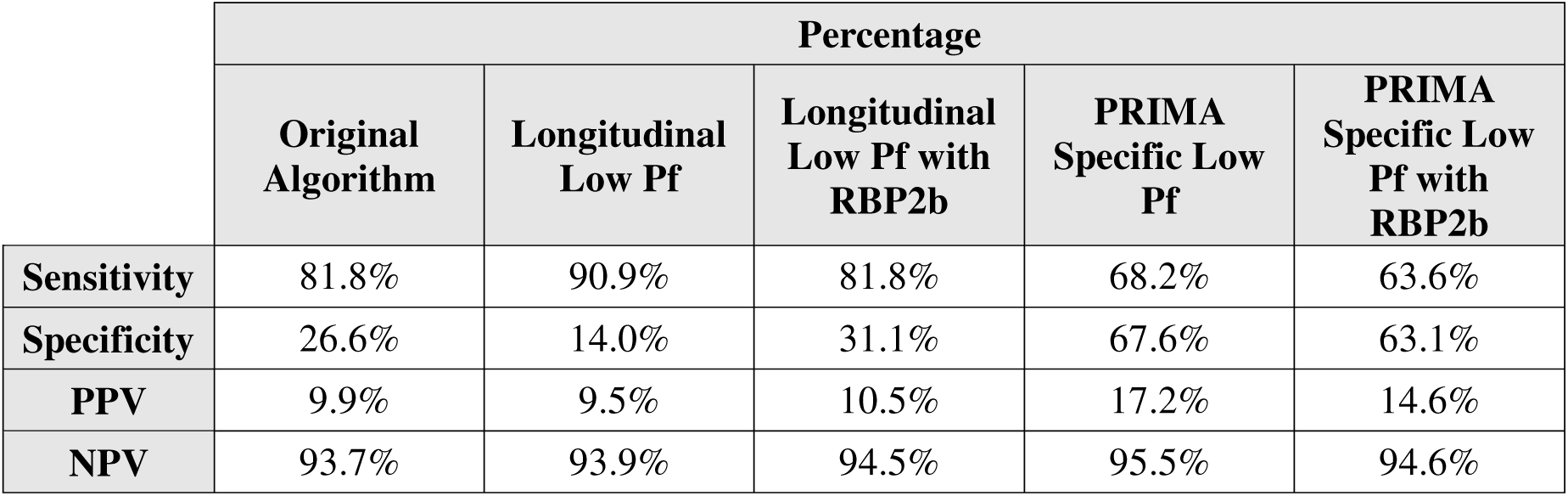
Comparison of Sensitivity, Specificity, PPV and PPV for Different Algorithms using maximised sensitivity & specificity. Longitudinal Low Pf excludes *P. vivax* proteins with known (low-level) cross-reactivity in *P. falciparum* patients; Longitudinal Low Pf with RBP2b is the same but with RBP2b added back in. PRIMA Specific Low Pf refers to an algorithm trained on the PRIMA data itself, using only the *P. vivax*-specific antigens. PRIMA Specific Low Pf with RBP2b is the same but with RBP2b added back in.

### Impact of past exposure and transmission intensity on *P. vivax* SEM classification performance’

The *P. vivax* SEMs have been designed for use in low endemic settings where malaria elimination efforts are targeting the hidden parasite reservoir often in asymptomatic individuals.^20^ When the *P. vivax* SEMs were tested in higher endemic settings, sensitivity and specificity declined. We hypothesized that the degree of past exposure present in areas with higher transmission intensity^15,20^, may have contributed to the poor specificity of the *P. vivax* SEMs in the patients enrolled into the PRIMA study with *P. falciparum* malaria. We explored this through the surrogate measures of the study site, age, and gender.

Study samples were collected from three sites^7^, Bangladesh (annual parasite index (API) 21.06), Indonesia (API in three study regions varying from 6.07-36.98)^7^ and Ethiopia (API unknown)^7^. The risk of *P. vivax* parasitaemia during follow-up varied across the three sites, ranging from 12.7% in Ethiopia, 16.9% in Bangladesh, and 2.2% in Indonesia.^7^ Although this variance may have been due to differences in local transmission intensity and risk of hypnozoite carriage, the different blood-stage drugs may have also played a significant role. In Indonesia patients were treated with dihydroartemisinin–piperaquine, a slowly eliminated drug, whereas in Ethiopia and Bangladesh, patients were treated with artemether–lumefantrine, which has a much shorter elimination and post treatment prophylaxis. A small number of events were observed in Indonesia (1 out of 50 patients) and Bangladesh (3 out of 24 patients), posing challenges in assessing classification performance per country; these are presented in the supplement (Figure S1). Due to the larger sample size, the results are driven by data from the patients enrolled in Ethiopia, however the sensitivity and specificity were consistent across countries.

IgG levels were compared across sites for the 8 *P. vivax* SEMs in the original algorithm (Figure 2A). No significant differences in antibody levels were observed across countries for Pvs16, MSP8, and RBP2b. In contrast, PTEX150 exhibited significant variation in antibody levels, with higher responses observed in Bangladesh compared to Ethiopia, and in Indonesia compared to Ethiopia, indicating lower antibody reactivity in the Ethiopian cohort. Similarly, for Pv-fam-a, antibody responses were significantly higher in Bangladesh compared to Indonesia and Ethiopia. A similar trend was observed for PvEBP, with Bangladesh displaying significantly elevated antibody levels relative to Indonesia. MSP5 followed this pattern, with significantly higher antibody responses in Bangladesh than in Indonesia. MSP1-19 antibody levels were significantly elevated in Ethiopia relative to both Indonesia and Bangladesh. Thus, whilst there were differences by antigen and site, the overall trend was towards higher IgG levels in Bangladesh compared to the other two countries, though notably no significant difference was observed for the antigen with the most importance in the classification algorithm – RBP2b.

**Figure 2.**
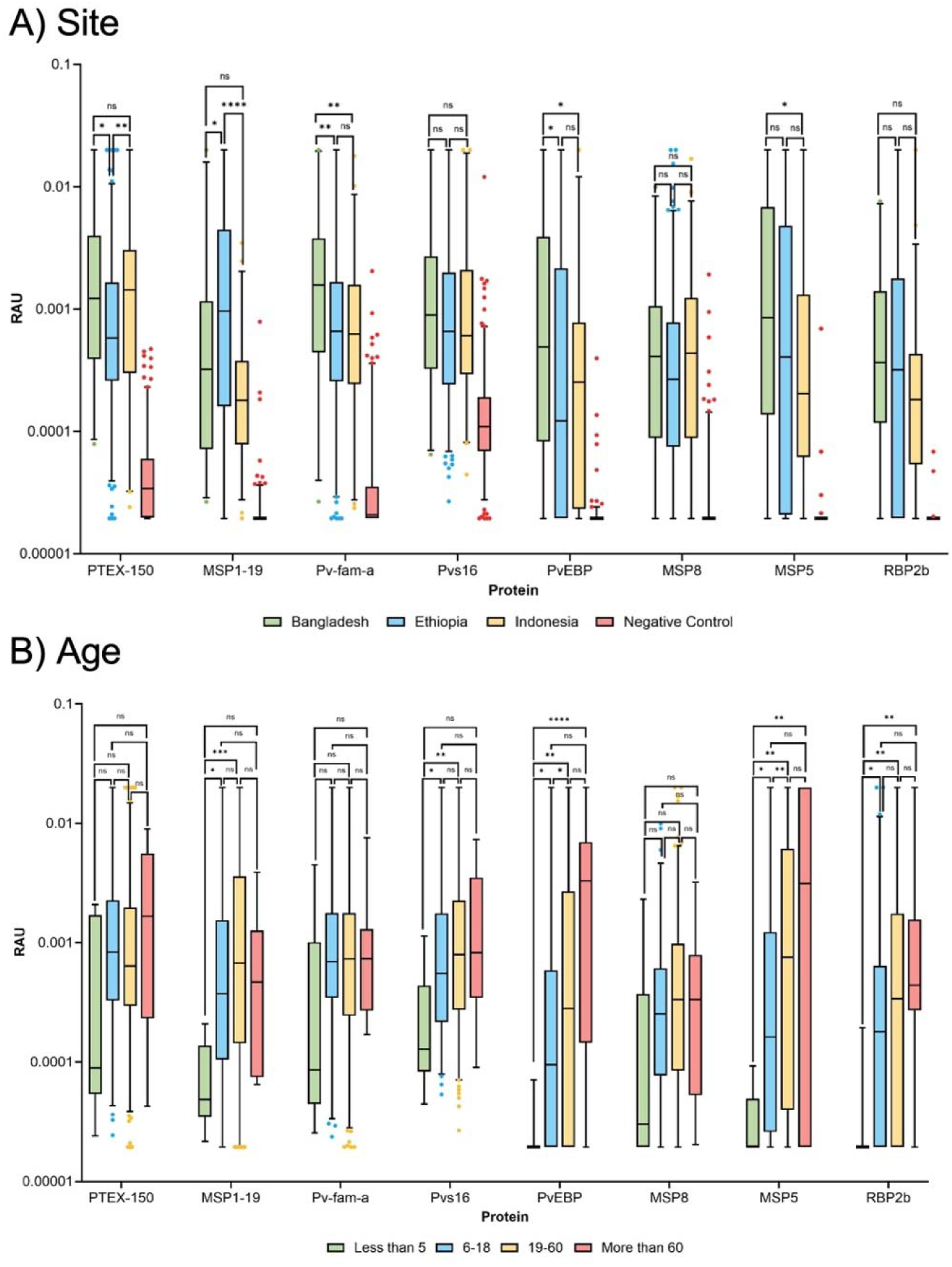
IgG antibody levels against the *P. vivax* SEM protein panel across study sites and by age group. The relative antibody units (RAU) were generated from median fluorescence intensity (MFI) values obtained using Luminex MAGPIX, then converted into RAU using a 5-parameter logistic regression model. Box plots represent the distribution of IgG levels for each protein, with (A) study sites indicated by color: Bangladesh (green, n=49), Ethiopia (blue, n=349), and Indonesia (yellow, n=99), while negative controls are shown in pink, or (B) with age groups indicated by colour: Less than 5 years (green, n=9), 6–18 years (blue, n=149), 19–60 years (yellow, n=305), and More than 60 years (red, n=11). Statistical comparisons were performed using the Kruskal-Wallis test then followed by the Dunn post-hoc test, with significance levels indicated as follows: * (p < 0.05), ** (p < 0.01), *** (p < 0.001), **** (p < 0.0001), and ns (no significant difference).

Age is a surrogate marker of exposure, with a clear trend for higher antibody levels in older individuals, signifying greater lifetime exposure. We categorised patients into four age groups: <5 years, 6–18 years, 19–60 years, and >60 years, noting that no children were enrolled in Ethiopia. The rationale for these age groupings is based on societal roles and life stages: the 6–18 age range represents the typical school-going population, 19–60 years represents the working-age population, and >60 years is classified as the elderly. Comparisons between these age groups demonstrated significant variations in antibody level for several proteins (6/8) (Figure 2B), with adults exhibiting higher levels than children. This trend was particularly clear for the antigens PvEBP, MSP5 and RBP2b.

In some settings, gender is considered a risk factor for exposure to *Plasmodium* spp. due to occupational exposure. For example, studies in Ethiopia and Bangladesh have reported that men of working age exhibit a higher immune response to *P. vivax*.^21,22^ However, in this study we observed no significant differences in IgG level between men and women for any of the 8 *P. vivax* SEMs (Figure S2).

### Improving classification accuracy of risk of *P. vivax* in *P. falciparum* patients

Investigations into the poor specificity of the *P. vivax* SEMs suggested that this was likely to be attributable to higher levels of prior exposure at the PRIMA study sites compared to sites where the *P. vivax* SEMs have been tested previously. We tested a different use of *P. vivax* SEMs; instead of relying on the algorithm trained in low-transmission settings, we developed a PRIMA study-specific algorithm tailored to the local epidemiology of malaria, using the serology data from the trial itself. To avoid overfitting, we used a 10-fold cross-validation, where the dataset was split into 10 folds, each serving once as a test dataset while the remaining 90% of the data was used for training. This process was repeated five times to ensure robustness. Proteins with potential *P. falciparum* cross-reactivity were excluded, and their removal did not reduce sensitivity in earlier analyses. The PRIMA-specific algorithm showed an improved balance between sensitivity and specificity, with a specificity of up to 68.2% (Table 2). However, this was still lower than the specificity of 80% reported in the original longitudinal cohort studies.^11^

**Table 2.** Performance Summary of Each Algorithm with Different Specificities Target.

|  | original algorithm |  |  |  | Longitudinal Low Pf |  |  |  | Longitudinal Low Pf with RBP2b |  |  |  | PRIMA Specific Low Pf |  |  |  | PRIMA Specific Low Pf with RBP2b |  |  |  |
| --- | --- | --- | --- | --- | --- | --- | --- | --- | --- | --- | --- | --- | --- | --- | --- | --- | --- | --- | --- | --- |
|  | Sensitivity | Specificity | NPV | PPV | Sensitivity | Specificity | NPV | PPV | Sensitivity | Specificity | NPV | PPV | Sensitivity | Specificity | NPV | PPV | Sensitivity | Specificity | NPV | PPV |
| **Maximised 80-80** | 81.8% | 26.6% | 93.7% | 9.9% | 90.9% | 14.0% | 93.9% | 9.5% | 81.8% | 31.1% | 94.5% | 10.5% | 68.2% | 67.6% | 95.5% | 17.2% | 63.6% | 63.1% | 94.6% | 14.6% |
| **Specificity 85%** | 77.3% | 34.2% | 93.8% | 10.4% | 81.8% | 36.5% | 95.3% | 11.3% | 72.7% | 41.9% | 93.9% | 11.0% | 36.4% | 86.5% | 93.2% | 21.1% | 36.4% | 86.0% | 93.2% | 20.5% |
| **Specificity 90%** | 72.7% | 47.7% | 94.6% | 12.1% | 77.3% | 45.5% | 95.3% | 12.3% | 59.1% | 54.5% | 93.1% | 11.4% | 27.3% | 89.2% | 92.5% | 20.0% | 22.7% | 89.6% | 92.1% | 17.9% |
| **Specificity 95%** | 45.5% | 68.5% | 92.7% | 12.5% | 54.5% | 61.7% | 93.2% | 12.4% | 31.8% | 76.1% | 91.8% | 11.7% | 22.7% | 94.1% | 92.5% | 27.8% | 18.2% | 94.6% | 92.1% | 25.0% |

To improve specificity, we adjusted the random forest voting thresholds to target specificities of 85%, 90%, and 95%. The results for each algorithm at these thresholds are presented in Table 4. As the specificity target increased, sensitivity decreased for all algorithms. For example, at an 85% specificity target, the “PRIMA-specific Low Pf” and “PRIMA-specific Low Pf with RBP2b” algorithms achieved specificities of 86%, with sensitivities of 36% and 34%, respectively. The modified specificity targets were also tested on the original and low Pf adjusted algorithms trained on the longitudinal cohort data, reaching a more balanced sensitivity and specificity at either the 90% or 95% specificity target, but with values much lower than the goal of 80% sensitivity and 80% specificity. The best-balanced result overall remained with the PRIMA specific low Pf algorithm at 68% sensitivity and 68% specificity (NPV was high with all options tested). These trade-offs between sensitivity and specificity are visualised through receiver operating curves (ROC) in Figure 3, with the area under the curve (AUC) used to quantify overall performance. The original model applied to the PRIMA dataset has an AUC of 0.583, indicating modest performance. The low Pf and low Pf with RBP2b models show slightly lower AUC values of 0.559 and 0.551, respectively. However, the dataset-specific models show significant improvement, with both the PRIMA-specific low Pf and PRIMA-specific low Pf with RBP2b models achieving an AUC of 0.716.

**Figure 3.**
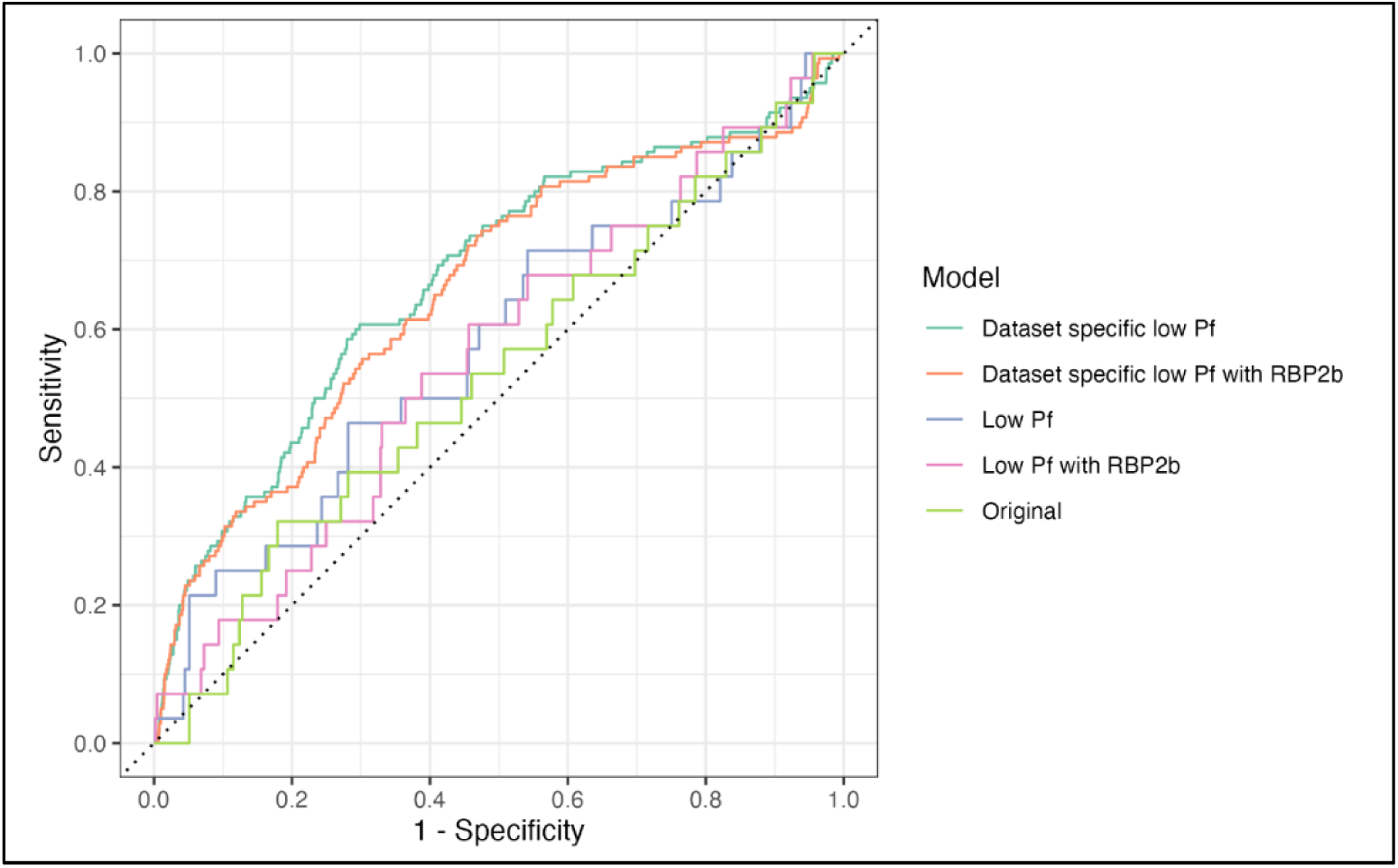
ROC curve of different algorithms tested on the PRIMA study samples. AUC values for each model: Original (0.583), Low Pf (0.559), Low Pf with RBP2b (0.551), Dataset specific low Pf (0.716), and Dataset specific low Pf with RBP2b (0.716).

## DISCUSSION

*P. vivax* continues to pose a key challenge for malaria elimination due to difficulties in its treatment^23^, control^24^ and surveillance^20^. Novel approaches need to be considered to eliminate the undetected hypnozoite reservoir that maintains transmission within communities. Universal radical cure is an approach that opportunistically eradicates *P. vivax* hypnozoites that may be present in the liver of patients presenting clinical *P. falciparum* malaria in co-endemic areas. The PRIMA study demonstrated that high-dose short-course primaquine was able to reduce the risk of subsequent *P. vivax* parasitaemia in *P. falciparum* patients within 63 days by five-fold, and this was most apparent in Ethiopia, where incidence of both species was greatest.^7^ Universal radical cure has potential to reduce the hypnozoite reservoir, however the public health benefit will likely vary significantly between different endemic settings.^25^ Universal radical cure is not yet recommended in WHO malaria treatment guidelines, and countries may be reluctant to introduce this approach. Concerns are predominantly focused on the risk-benefit ratio since patients who do not harbour hypnozoites are potentially exposed to 8-aminoquinoline treatment. The latter can be reduced by G6PD testing, although this is associated with logistical and financial challenges. A more targeted approach to universal radical cure using *P. vivax* SEMs as an additional screening tool could alleviate some of those concerns. Whilst we found limited specificity of *P. vivax* SEMs in patients with *P. falciparum* malaria residing in these endemic settings, the NPV was high. Thus, *P. vivax* SEMs could be considered to limit exposing people to 8-aminoquinolines unnecessarily, and to reduce the numbers of G6PD-tests and treatment doses required for unrestricted universal radical cure. Likewise, the sensitivity was high (82%), suggesting that most individuals with a subsequent episode of *P. vivax* were identified. Whether this is high enough to result in the reduction of transmission, and whether the costs of the *P. vivax* SEM assay are balanced by reduced numbers of G6PD-tests and 8-aminolquinoline treatment doses, warrants for study.

We explored various reasons for the low specificity (27% when using the original algorithm), which was due to a high level of the *P. falciparum* patients being classified as sero-positive against the *P. vivax* SEMs but who did not have subsequent *P. vivax* parasitaemia detected during follow-up. We modified the classification algorithm to remove potentially cross-reactive antigens (based on data from unpublished work). This provided no improvement in specificity, suggesting a different reason must be responsible for the high seropositivity rates. This may warrant further research, as our antigen selection was based on the cross-reactivity of *P. vivax* antigens and their *P. falciparum* orthologs using data from low-transmission settings; antibody cross-reactivity between these orthologs may differ in areas of higher transmission. This is broadly important for the use of the *P. vivax* SEMs for surveillance and guiding interventions in areas co-endemic for both species, for example within *P. vivax*-endemic regions of Africa (i.e. Madagascar, Ethiopia).

The high levels of seropositivity and low specificity of the algorithm are likely due to the PRIMA study sites which were in areas of higher transmission settings than previously studied (at least for Ethiopia and Bangladesh). The *P. vivax* SEMs were designed for use in low-transmission settings that are approaching elimination. In higher transmission settings, antibody responses to the same *P. vivax* antigens can last for longer^26^, and *P. vivax* SEM performance will be lower.^27^ The study sites in Bangladesh, Ethiopia and Indonesia were selected because of relatively high transmission rates (in the context of their country-specific epidemiology) to facilitate recruitment of clinical malaria patients. We developed an algorithm trained on the PRIMA serological data itself where specificity was improved to 68% (with reduced sensitivity to 68%), though this would ideally be validated on an independent cohort to test its generalisability in similar transmission settings. Other factors may have contributed to the poor performance of the *P. vivax* SEMs in *P. falciparum* patients in these settings. Patient follow-up was curtailed at 63 days and whilst this may have been sufficient to detect most early relapses ^28^, it will not have identified all individuals with later relapses. *P. vivax* relapses can occur at least up to 9-months following the primary infection^16^, thus some individuals identified as being sero-positive may have had an episode of *P. vivax* after the end of follow up. Furthermore, screening for *P. vivax* parasitaemia was done by microscopy and not by PCR, and low-density sub-microscopic infections may have been missed.^29^ It is possible that some of the sero-positive individuals may have had subsequent *P. vivax* infections that were missed by microscopy.

## CONCLUSIONS

Overall, our data provides limited evidence for the use of *P. vivax* SEMs to guide radical cure of patients with *P. falciparum*. The high sensitivity and a high NPV of the testing offer potential benefits in reducing the numbers of individuals who need to be G6PD-screened and exposed to radical cure compared to a universal treatment of all patients presenting with *P. falciparum*. However, this needs to be balanced against the increased cost and logistical considerations. The usefulness of *P. vivax* SEMs likely depends on how countries and program managers view the cost-benefit ratio of these options. Our study was conducted in relatively high transmission settings, with most data from Ethiopia; hence it is possible that the *P. vivax* SEMs may be more accurate in guiding radical cure in *P. falciparum* patients in lower transmission settings (more similar to where the algorithm was trained). This may result in better performance of the SEMs, however the risk of *P. vivax* following *P. falciparum* infection decreases with reduced overall disease burden^30^, likely limiting the impact of universal radical cure on transmission. In these settings, active community-wide approaches such as *P. vivax* serological testing and treatment (PvSeroTAT)^20^ may be more beneficial. Importantly, the SEM assay described requires a laboratory setting and trained staff, however, it is feasible to develop a point-of-contact test to measure these antibody biomarkers. The original classification algorithm is currently available as a RShiny App and is highly usable.

## Data Availability

All data produced in the present work are contained in the manuscript (supplemental data file provided).

## ACKNOWLEDGMENTS

We acknowledge the efforts of all members of the original study teams who collected the PRIMA samples, along with all study participants. We thank the following organisations for the donation of malaria-naïve control plasma: the Australian and Thai Red Cross (the latter with support from Jetsumon Sattabongkot), the Rio de Janeiro State Blood Bank (with support from Andre M. Siqueira), and the Volunteer Biospecimen Registry at WEHI. We acknowledge Kenneth Wu for calculating the sequence identity data presented in Table S1, as part of another unpublished project, and for sharing data that guided exclusion of *P. falciparum* cross-reactive antigens. We thank Julie Healer for assistance with ethical approvals and sample shipments. We acknowledge proteins provided by Wai-Hong Tham (WEHI) and proteins produced and purchased from ZiP Diagnostics and the WEHI Protein Production Facility (Marija Dramicanin). We thank Anju Abraham, Macie Lamont and Ramin Mazhari for laboratory support and assistance. We also acknowledge the Victorian State Government Operational Infrastructure Support and Australian Government NHMRC IRIISS.

## AUTHOR CONTRIBUTIONS

Conceptualization: IM, KT, RJL. Data curation: MH. Formal analysis: MH, LS, DA. Resources: TSD, MC, MSA, MSH, JKB, IS, ARA, RN, RP, IM, KT, RJL. Supervision: KT, RJL, LS. Visualisation: MH, LS, DA. Writing – original draft: MH, KT, RJL. Writing – reviewing & editing – all authors. MH, LS and DA performed statistical analyses. MH, LS and RJL had unrestricted access to the data. All authors agree to submit the manuscript, read, and approved the final draft and take full responsibility of its content, including the accuracy of the data and the fidelity of its statistical analysis.

