## Supplemental Tables and Figures for "Evaluating the Performance of Predicting *Plasmodium vivax* Infection Risk Using Serological Markers in Patients with *Plasmodium falciparum* Malaria"

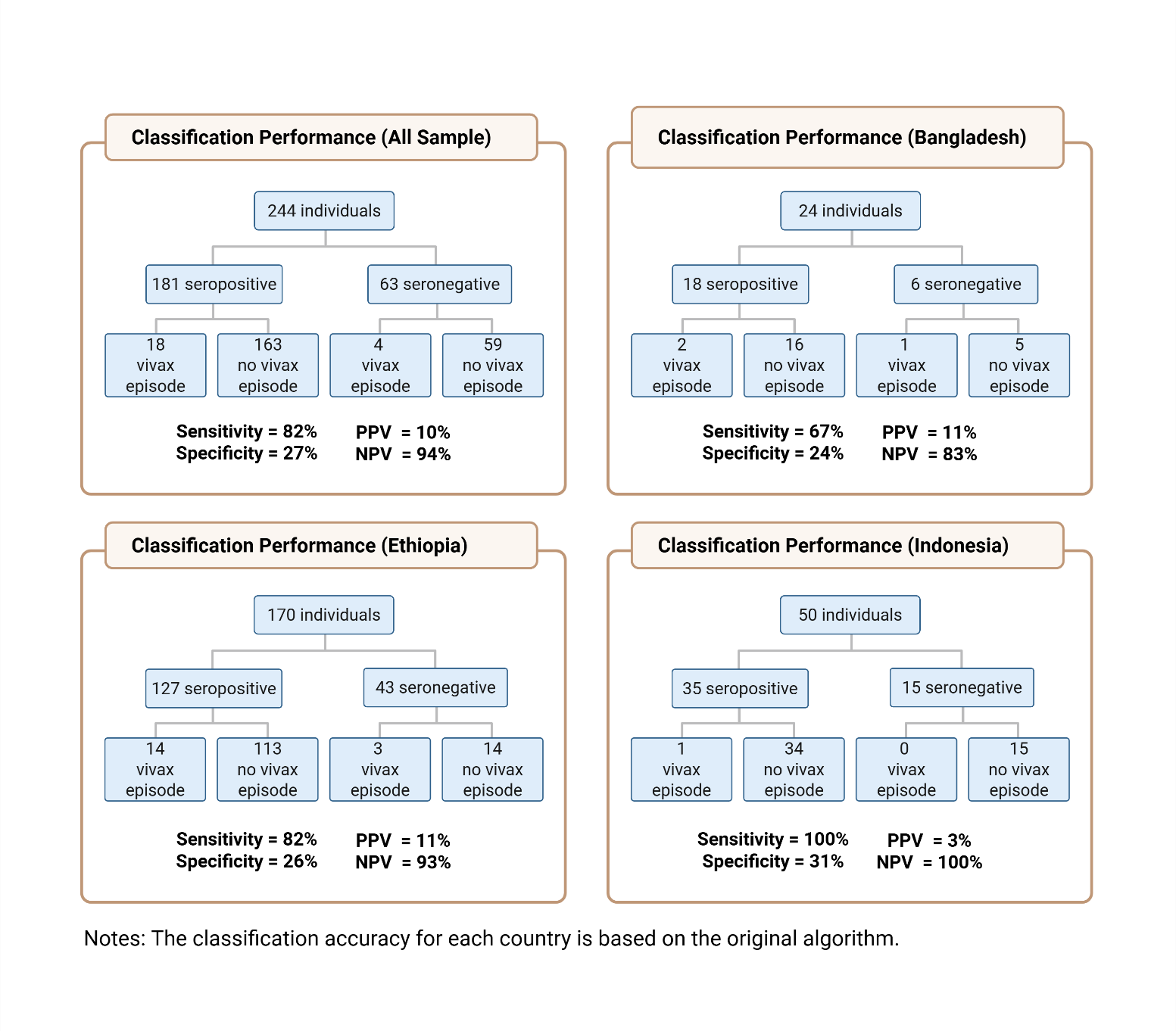

**Figure S1.** Classification performance breakdown by country. Classification is via the original algorithm only.

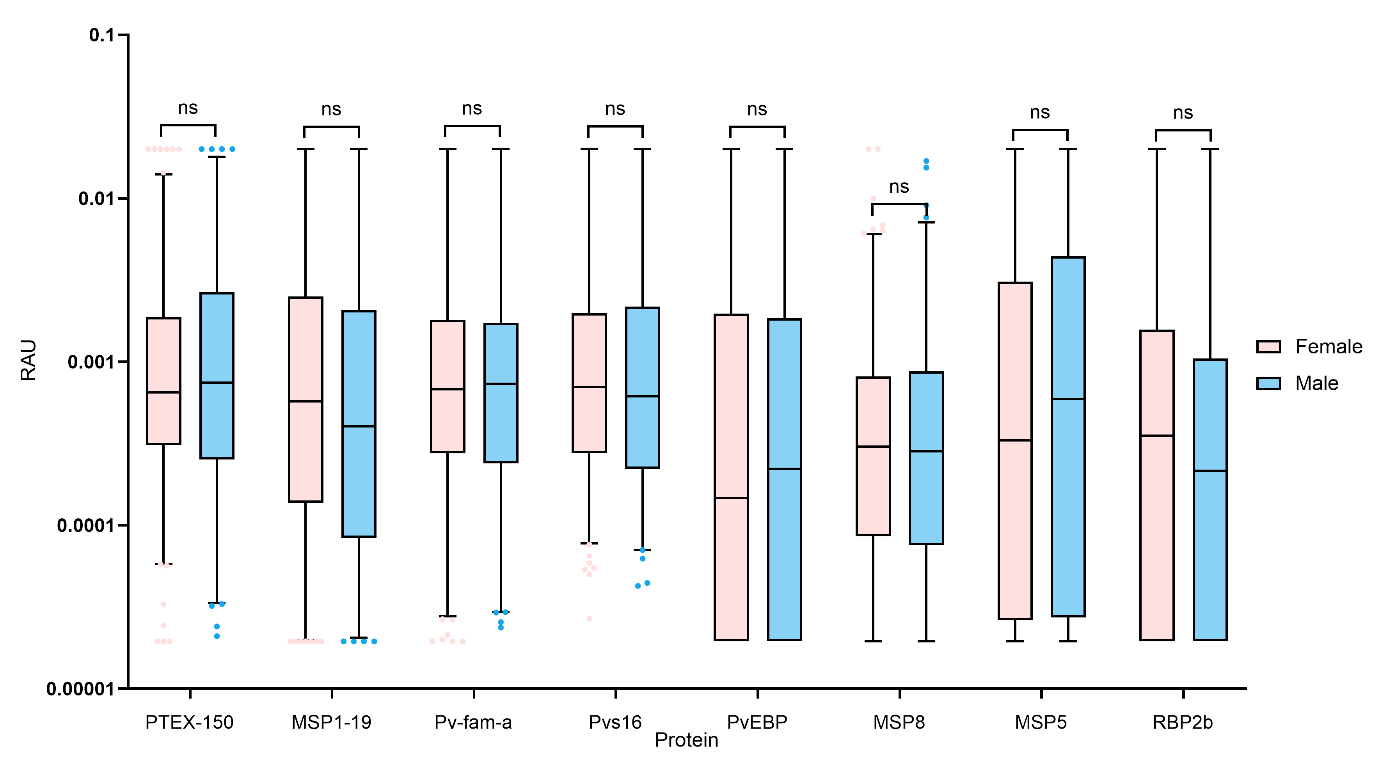

**Figure S2.** The RAU were derived from MFI values obtained using Luminex MAGPIX, then converted into RAU using a 5-parameter logistic regression model. Box plots represent the distribution of IgG levels for each protein, with gender by colour: Female (pink), and Male (blue). Statistical comparisons between gender were performed using the kruskall-wallis test, with significance level indicated as follows: * (p < 0.05), ** (p < 0.01), *** (p < 0.001), **** (p < 0.0001), and ns (no significant difference). Sample sizes: Male (n=177), Female (n=297).

**Table S1.** Algorithms Used in this Study

| **Algorithm Name** | **Notes** |
| --- | --- |
| Original Algorithm | Base algorithm trained from the longitudinal cohort |
| Low Pf Algorithm | Base algorithm trained from the longitudinal cohort, excluding protein with high reactivity with *P. falciparum* |
| Low Pf Algorithm with RBP2b | Base algorithm trained from the longitudinal cohort, excluding protein with high reactivity with *P. falciparum* but still including RBP2b |
| Dataset Specific Low Pf | Algorithm trained from the PRIMA dataset excluding protein with high reactivity with *P. falciparum* |
| Dataset Specific Low Pf with RBP2b | Algorithm trained from the PRIMA dataset excluding protein with high reactivity with *P. falciparum* but still including RBP2b |

**Table S2.** Confusion Matrix

|  |  | **Classification** | |
| --- | --- | --- | --- |
|  |  | **Seropositive** | **Seronegative** |
| **Actual Condition** | **Relapse** | True Positive | False Negative |
|  | **No relapse** | False Positive | True Negative |

**Table S3.** Bead Region and P. vivax Protein List

| **Bead Region** | **Protein Name** | **Antigen ID** | **Amount (μg)** | **Construct, aa (size)** |
| --- | --- | --- | --- | --- |
| 54 | LF-005 (Pv-fam-a) | PVX_096995 | 1.5 | 61-end (420) |
| 57 | LF-010 (MSP5) | PVX_003770 | 0.1 | 23-365 (343) |
| 34 | LF-016 (MSP1-19) | PVX_099980 | 2.0 | 1622-1729 (108) |
| 53 | MSP8 | PVX_097625 | 2.0 | 24-463 (440) |
| 61 | EBP-II (PvEBP) | KMZ83376.1b | 0.3 | 109-432 (324) |
| 51 | RBP2b (P87) | PVX_094255 | 0.15 | 161-1454 (1294) |
| 15 | PTEX-150 | PVX_084720 | 0.5 | 24-908 (885) |
| 25 | Pvs-16 | PVX_000930 | 50 | 31-end (110) |
